# Pharmacokinetic-pharmacodynamic analysis of vismodegib in patients with basal cell carcinoma (the OPTIVISMO study)

**DOI:** 10.1101/2025.02.28.23295007

**Authors:** Pierre Mora, Sabrina Ayadi, Benjamin Sourisseau, Fabien Xuereb, Marie Beylot-Barry, Sarah Djabarouti

**Affiliations:** University Hospital Bordeaux and INSERM U1034; University Hospital Bordeaux; University Hospital Bordeaux and Bordeaux Institute of Oncology (BRIC)

**Keywords:** Basal cell carcinoma, chromatography–tandem mass spectrometry (HPLC–MS/MS), adverse effects, pharmacokinetic – pharmacodynamics, vismodegib

## Abstract

**WHAT IS KNOWN AND OBJECTIVE:** Vismodegib is indicated in patients with symptomatic metastatic basal cell carcinoma (BCC) or locally advanced BCC when surgery or radiotherapy are not appropriate. The significant efficacy of vismodegib in responding patients and lack of therapeutic alternatives are counterbalanced by intolerance and severe adverse events (AEs), leading to discontinuation in 30% of patients. We aimed to evaluate the relationship between vismodegib pharmacokinetics (PK) and occurrence of AEs, and to investigate the association between concentrations and response in terms of efficacy.

**RESULTS:** Mean of trough vismodegib plasma concentrations ranged from 3.9 mg/L to 30 mg/L per patient, with an overall mean of 11.8 (± 5) mg/L. A high correlation between total vismodegib and alpha-1 acid glycoprotein (AAG) levels was observed (Spearman’s ρ = 0.6733, p-value =1.662e-12). Inter-individual variability was significant (CV% of 42%).

Patients with stable and progressive disease had a significantly higher median vismodegib plasma concentration than those with partial and complete response (p = 0.03).

Tumor volume ranged from 0 to 12 292 135 mm^3^ in our cohort. Mean tumor volume slope was -1187.97 (± 9734).

**WHAT IS KNEW AND CONCLUSION:** We explored PK/pharmacodynamic (PD) relationships of vismodegib in patients with BCC. This is the first study which reported PK data obtained in BCC patients treated with vismodegib in a real-life clinical practice.

Our study confirmed the strong influence of AAG levels on vismodegib protein binding. Concerning the PK/PD relationship evaluation, we notably observed that patients with the lowest plasma concentrations respond best to treatment. Our mathematical estimation of tumor volume showed that between the beginning and the end of the study, tumor growth was positively correlated to vismodegib levels, which was in line with the correlation observed for efficacy/safety data.

## 1 WHAT IS KNOWN AND OBJECTIVES

Vismodegib is a Hedgehog pathway inhibitor indicated for the treatment of adults with metastatic basal cell carcinoma (BCC), or locally advanced BCC that has recurred following surgery or in patients who are not candidates for surgery or radiation. It was first approved by the FDA in 2012 and EMA in 2013 based on outcomes from the ERIVANCE study [1–3].

The pharmacokinetic profile of vismodegib is characterized by nonlinearity, high affinity, and saturable plasma protein binding to alpha-1 acid glycoprotein (AAG) with total vismodegib concentrations correlating closely to AAG concentrations, as well as low affinity binding to serum albumin [4–6].

At the standard dose of 150 mg/day orally, vismodegib is associated with many AEs, such as cramps, alopecia, dysgeusia, weight loss and others AEs observed in clinical practice, compromising compliance, and often leading to treatment discontinuation [7].

The OPTIVISMO clinical trial is a prospective, single-center, descriptive cohort study conducted by the hospital-university pharmacy team and the dermatology oncology Department, in collaboration with the Department of pharmacology (Bordeaux University Hospital). In our clinical practice, we observed that more than 30% of patients stopped treatment prematurely due to the appearance of AEs. We hypothesized that the occurrence of these adverse effects would be linked to high plasma concentrations of vismodegib. To date, vismodegib pharmacokinetic data are scarce and only available from controlled trials. To our knowledge, this is the first study that explored the pharmacokinetics of vismodegib in a context of a real-life clinical use.

The objective of the OPTIVISMO study was to assess the relationship between plasma concentrations of vismodegib, and the occurrence of adverse effects within 6 months of inclusion in the study. We also aimed to demonstrate a relationship between a patient’s exposure to vismodegib and a given efficacy/tolerance profile.

## 2 METHODS

### 2.1 Research description

The OPTIVISMO study is a single-center cohort-type study with prospective collection of safety and efficacy data, and evaluation of their relationship with total form of vismodegib plasma concentrations in patients with metastatic BCC, or advanced BCC. Patients were followed for 6 months. The CHU of Bordeaux is the promoter of this study. The first patient was enrolled on September 3, 2018. The enrollment period was 18 months, and the end of patient recruitment was May 12, 2021. All patients had signed their informed, free, and written consent, and the study was approved by the local Ethics Committee.

### 2.2 Study cohort

Twenty-seven patients were enrolled. The inclusion criteria were: (i) age ≥ 18 years; (ii) patients with BCC or histologically proven adnexal carcinoma who have started or have been on vismodegib for at least 21 days. The exclusion criteria were patients: (i) who have discontinued treatment due to non-response or progression; (ii) with confusional syndrome; (iii) with a contraindication to vismodegib; (vi) who were pregnant or lactating.

### 2.3 Research strategies and procedures

Vismodegib was given in 150 mg capsules daily until disease progression or unacceptable toxicity (study does not change practice). The patients were seen in consultation every month to collect clinical-biological and anthropometric data. The pharmacokinetic protocol provided for two blood samples to be taken for each patient just before drug intake, respectively for vismodegib and AAG level measurement. Blood was drawn when the pharmacokinetic steady-state was reached i.e., at least after 21 days of treatment, to assess trough concentration.

Safety data were collected according to the NCI-CTCAE criteria (Common Terminology Criteria for Adverse Events version 5.0 of the National Cancer Institute) and classified by grade according to the severity of the AEs [8]. The clinical response in terms of efficacy was evaluated and classified into 4 categories (progression, stability, partial response, and complete response) according to a standard for measuring tumors, RECIST (response evaluation criteria in solid tumors) guidelines (Figure1) [9]. ECOG (Eastern Oncology Cooperative Group) performance status was also noted. The performance scale measured the impact of illness on a patient’s abilities, and was made up of 6 grades, ranging from no impact to patient death [10].

### 2.4 Sample processing

For total vismodegib determination in plasma, 10 μL of plasma were diluted with 500 μL of 5% formic acid in water solution and 20 μL of internal standard, and the extracted by a solid phase extraction (SPE) process. Briefly, the extraction plate was rehydrated with 1000 μL of methanol and then conditioned with 1000 μL of water-5% formic acid. The diluted samples were loaded to the SPE wells. Then, they were washed with 1000 μL of methanol and finally each well was eluted three times with 300 μL of 5% ammonia methanol solution. The eluate was then evaporated to dryness in a rotary concentrator under vacuum at 40°C and then reconstituted each with 100 μL of 1% formic acid in acetonitrile: water (80:20, v:v) and mixed for 5 min. Next, 2 μL of sample was injected into the high-performance liquid chromatography tandem mass spectrometry (HPLC–MS/MS) system.

### 2.5 Measurement methods

Total vismodegib concentrations in patient’s plasma were determined using the HPLC method developed and validated in the Department of pharmacology (Bordeaux University Hospital). Calibration standards ranged from 0.5 to 30 mg/L for plasma. The correlation coefficient of the validation range was greater than 0.99, and revealed an excellent linearity. Intra- and inter-day precision of the analytical assay were <15.0%, respectively. Intra- and inter-day accuracy ranged from 85.0% to 115.0%.

### 2.6 Pharmacokinetic analysis

We assessed the inter- and intra-individual variability of trough plasma concentrations of the total form of vismodegib by determining coefficients of variation (CV%), and then assessed the relationship between plasma levels of AAG and plasma concentrations of vismodegib. Safety and efficacy data were evaluated to investigate a possible relationship between vismodegib plasma concentrations and therapeutic response (tolerance/efficacy).

### 2.7. Pharmacodynamic analysis

For each patient, the tumor size was measured at each visit (size of the long axis and the short axis), and the tumor volume was estimated using the mathematical formula for volume estimation as follows: 4/3*π*x*y*z, where x, y and z represent diameters in the x, y, and z axes. Tumor volumes were calculated by substituting the x, y, and z diameters with linear dimensions to obtain a hypothetical ellipsoid 4/3*π*x²*y and expressed in mm^3^ [11]. Then, the longitudinal tumor volume slope was calculated for each patient using the last and the first available volume.

### 2.8 Statistical analysis

Pharmacokinetic data and pharmacodynamics values (tumor volumes) were expressed as mean (± standard deviation), and/or median [interquartile ranges, Q1; Q3]. Correlation between pharmacokinetic data and covariates (AAG level and other patient data), and between pharmacodynamic and pharmacokinetic data were established using the Spearman test. The Mann-Whitney test was used for comparison between groups. Tests were considered significant when the p-value was <0.05.

## 3 RESULTS AND DISCUSSION

### 3.1 Patients demographics

In the OPTIVISMO study, 27 patients were included. The gender distribution in the study was homogeneous with a male to female ratio, sex ratio, at 1.25. We noted that the average age was high, 72.9 years, with only three patients under 60 (Table 1). Two patients were finally withdrawn from the study because they refused to take vismodegib.

**TABLE 1.**
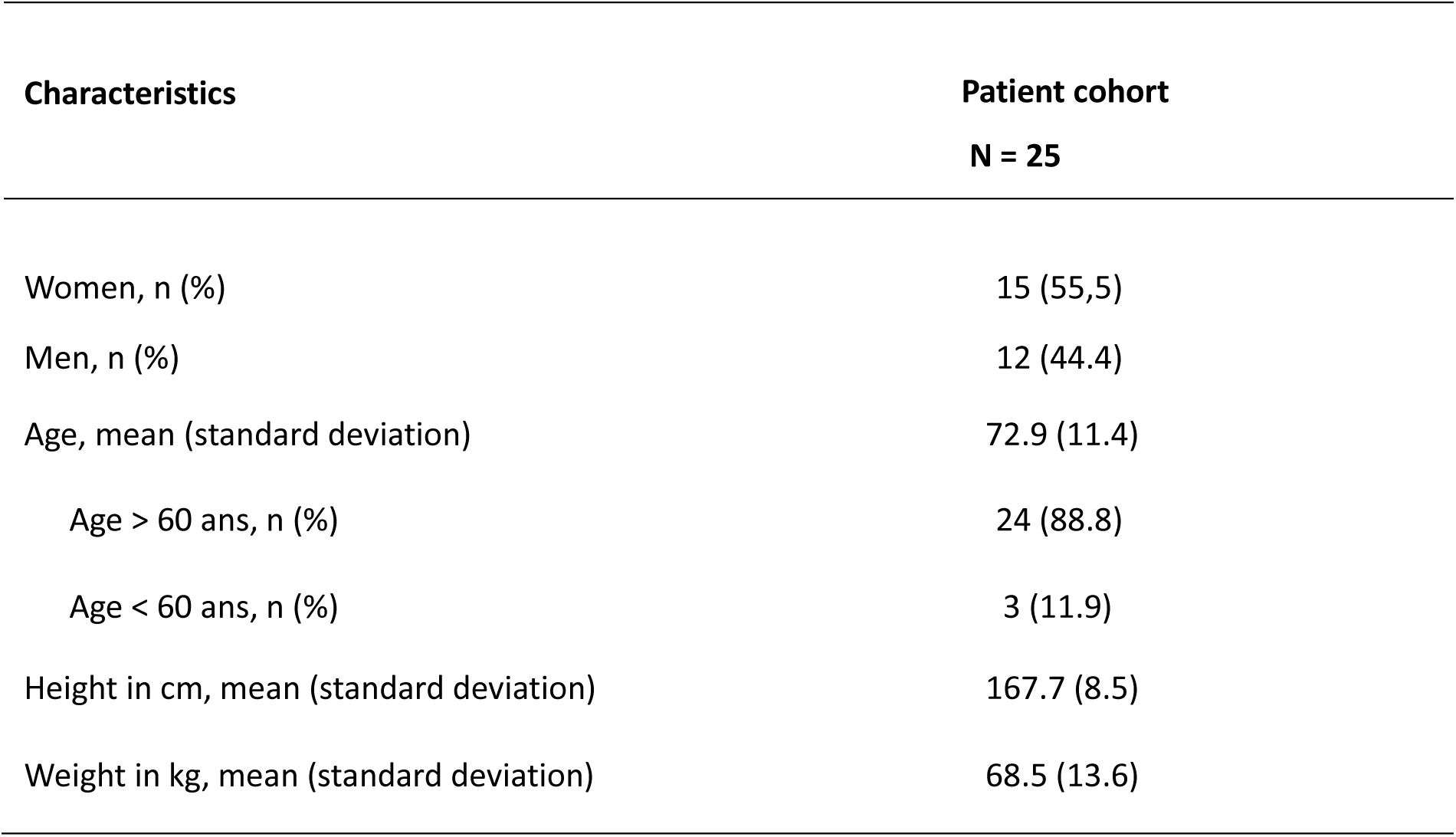
Summary of demographic and anthropometric characteristics of the patients included in the study.

| Characteristics | Patient cohort<br>N = 25 |
| --- | --- |
| Women, n (%) | 15 (55,5) |
| Men, n (%) | 12 (44.4) |
| Age, mean (standard deviation) | 72.9 (11.4) |
| Age > 60 ans, n (%) | 24 (88.8) |
| Age < 60 ans, n (%) | 3 (11.9) |
| Height in cm, mean (standard deviation) | 167.7 (8.5) |
| Weight in kg, mean (standard deviation) | 68.5 (13.6) |

### 3.2 Patients clinical characteristics

A total of 25 patients were followed during this study. The mean albumin level was 42.2 (± 4.3) g/L (range, 31.6 – 44.7), mean of creatinine (Cr) level was 77 (± 15.0) µmol/L (range, 48.5 – 124.2) and AAG mean was 0.8 g/L. The average duration of vismodegib was 5.8 (± 2.7) months (range, 2 – 3). The clinical characteristics of patients are detailed in Table 2.

**TABLE 2.**
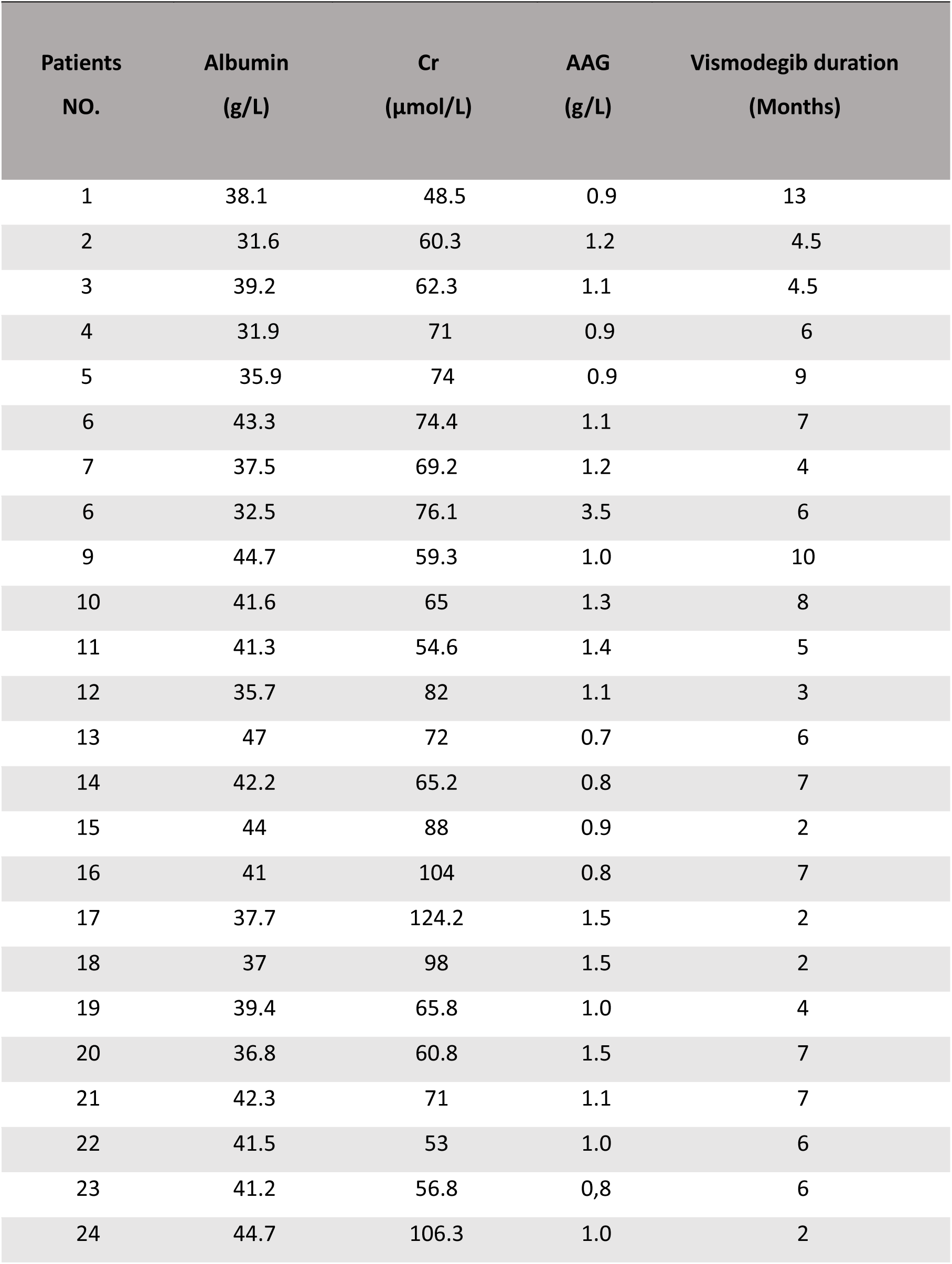
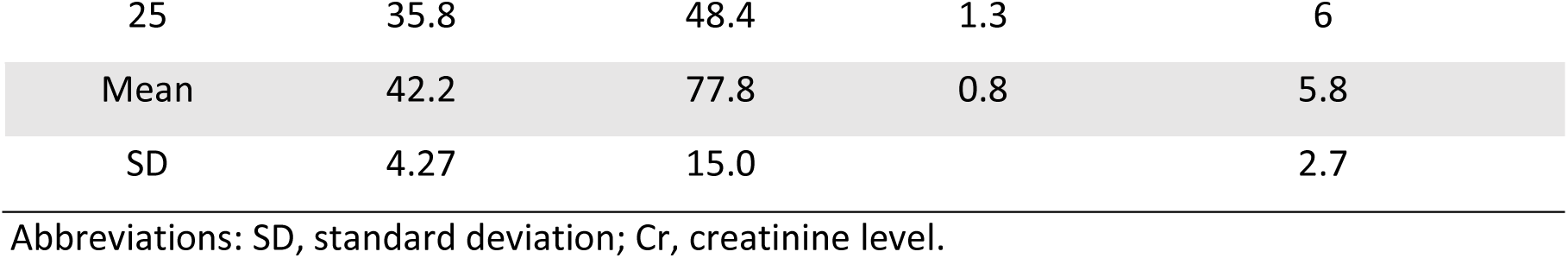
Basic characteristics of the 25 patients in the study.

### 3.3 Disease characteristics at inclusion

At inclusion, 11 patients were not taking vismodegib at the standard dosage of 150 mg once daily. The others had been taking vismodegib from 1 month for the most recent to 8 months for the oldest.

At the end of the study, 6 patients stopped taking vismodegib, and 6 other patients took vismodegib on a sequential schedule. Only 3 patients had an ECOG score greater than or equal to 2 (Table 3).

**TABLE 3.** Disease characteristics of patients at baseline.

|  | All patients<br>N= 25 |
| --- | --- |
| ECOG PS |  |
| Grade 0 | 13 (52) |
| Grade 1 | 9 (36) |
| Grade $\geq 2$ | 3 (12) |
| Lesions |  |
| Measurable lesions | 17 (68) |
| Non measurable lesions | 8 (32) |

At baseline exam, 17 patients had measurable lesions and 8 patients had non-measurable lesions.

### 3.4 Pharmacokinetic data

Mean residual vismodegib plasma concentrations per patient ranged from 3.9 mg/L to 30 mg/L with a mean of 11.8 (± 5) mg/L. The inter-individual variability was significant in our cohort with a CV% of 42 %. The intra-individual variability of vismodegib plasma concentrations at steady-state was low for 14 patients with a CV%< 30%, and high for 4 patients with a CV% greater than 50%. Vismodegib concentrations of patients measured during the follow-up are presented in the Figure 2.

**Figure 1.**
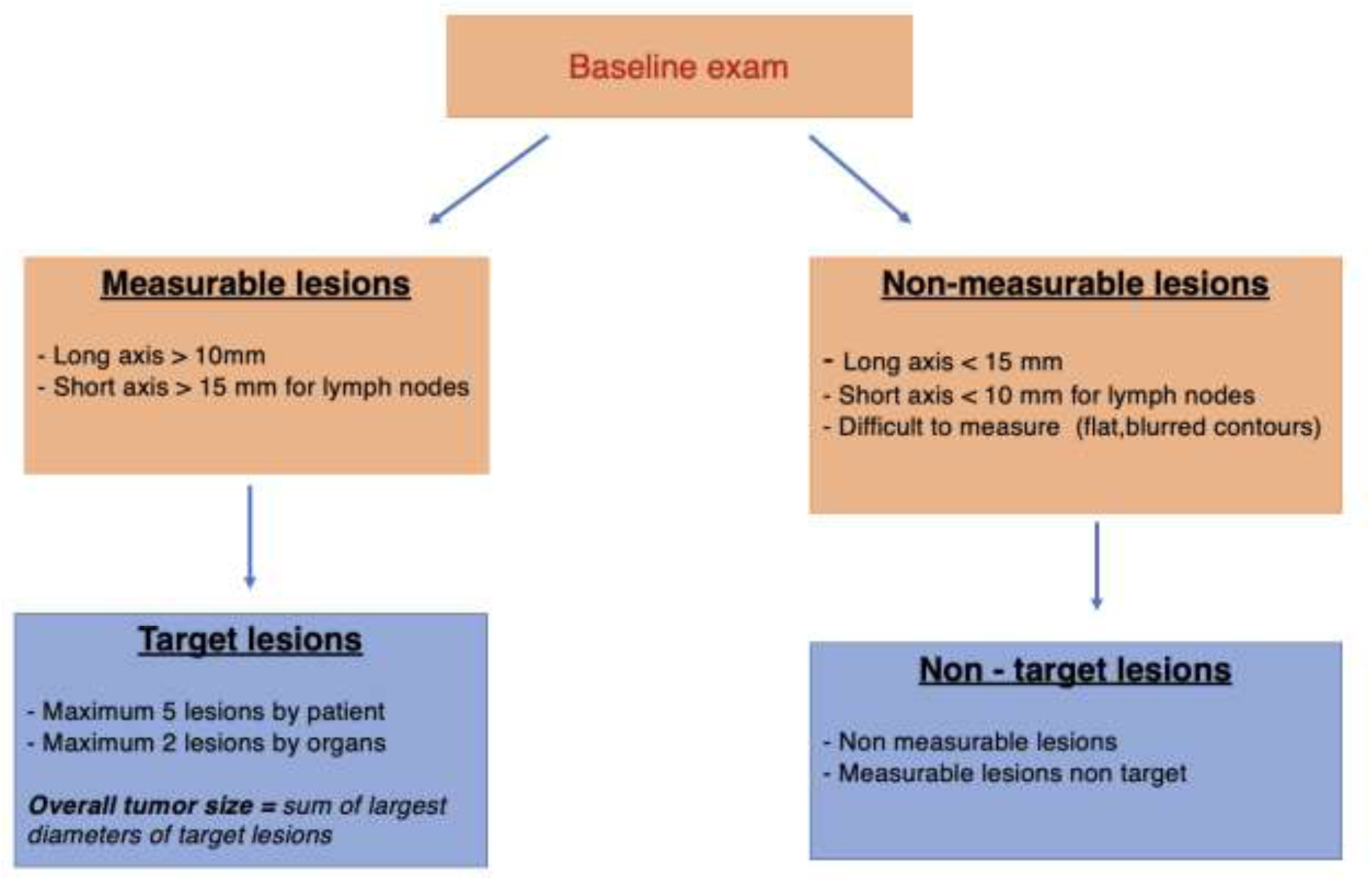
RECIST guidelines summary version (1.1).

**Figure 2.**
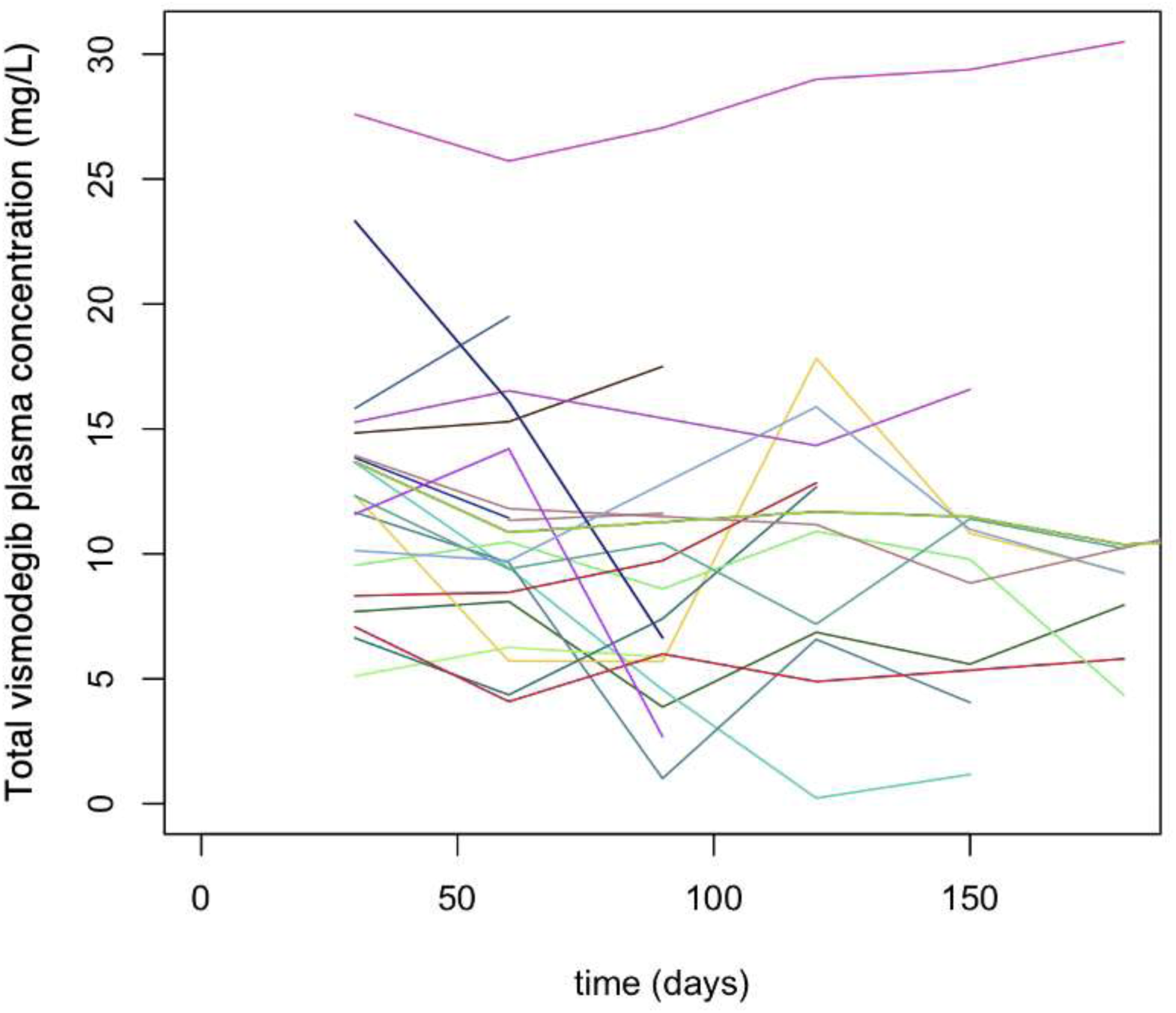
Total vismodegib concentrations of the patients of the OPTIVISMO study.

### 3.5. Pharmacokinetic-efficacy data relationship

We classified patients according to the best response obtained: on the one hand, patients with stable or progressing disease, and on the other, patients with partial and complete response disease.

Patients with stable (S) and progressive (P) disease had a significantly higher median trough vismodegib plasma concentration than patients with partial (PR) and complete (CR) response (according to the Mann-Whitney test, p = 0.03) (Table 4 and Figure 3). Other criteria differentiated these 2 groups, including a higher mean clearance (Mann-Whitney test, p=0.0095) and mean weight (Mann-Whitney test, p=0.0057), as well as lower mean age (Mann-Whitney test, p<0.0001) in patients with partial and complete response.

**Figure 3.**
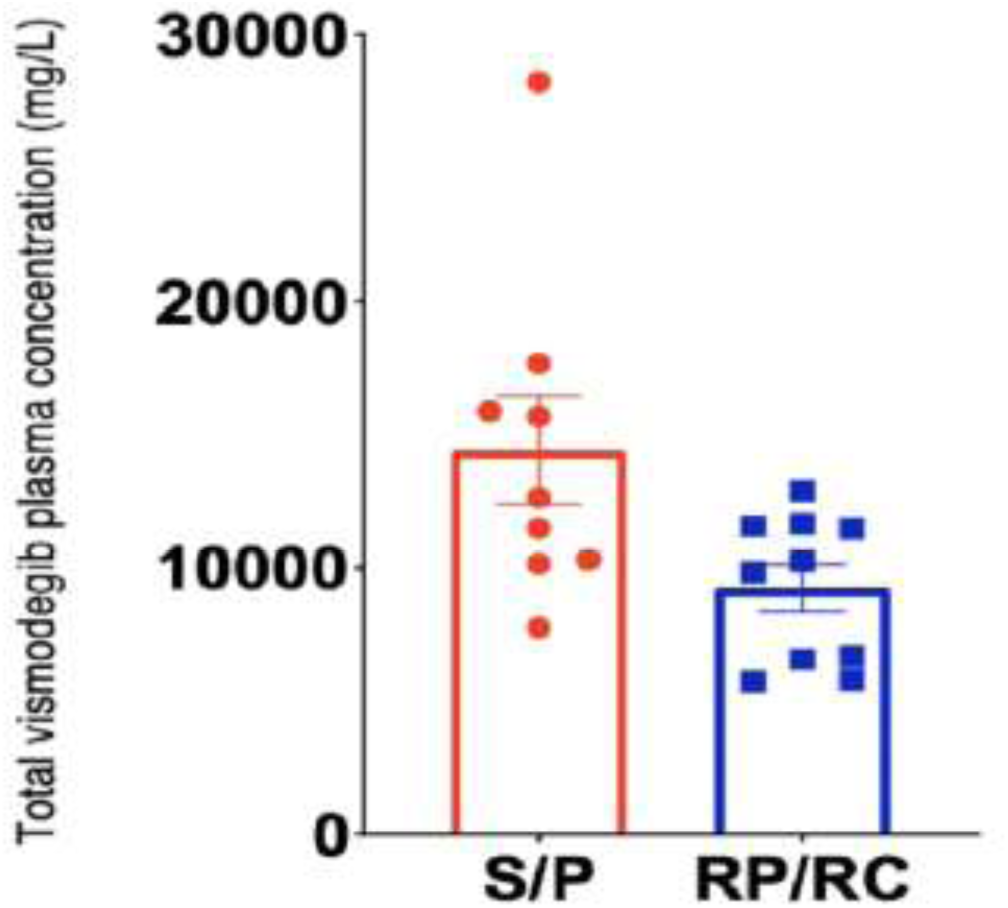
Box plots for clinical response according to RECIST criteria. Patients with stable (S) and progressive (P) disease had a significantly higher median trough vismodegib plasma concentration than patients with partial (PR) and complete (CR) response (p = 0.03)

**TABLE 4.** Median of trough vismodegib concentrations according to patient response.

| Patients responses | Median vismodegib concentrations (mg/L)<br>[Interquartile range] |
| --- | --- |
| RP/RC | 10 [11.2 ; 14.7] |
| S/P | 12 [7.3; 11.6] |

### 3.6. Pharmacokinetic-safety data relationship

All our patients experienced AEs. They were divided into 2 groups: grade 1 AEs and AEs of grade greater than or equal to 2. A higher trough concentration was observed in patients with grade 1 AEs; however, this result was not statistically significant (according to the Mann-Whitney test, p = 0.17) (Table 5).

**TABLE 5.** Median of trough vismodegib concentrations according to AEs grade.

| AEs grade | Median vismodegib concentrations (mg/L)<br>[Interquartile range] |
| --- | --- |
| Grade = 1 (n=26) | 11.8 [9.2 ; 14.7] |
| Grade $\geq 2$ (n=67) | 10.5 [8 ; 13.5] |

### 3 .7 Pharmacodynamic analysis

Tumor volume ranged from 0 to 12 292 135 mm^3^ in the OPTVISMO cohort. Mean tumor volume slope was -1187.97 (± 9734), indicating a global reduction of tumor size for patients under vismodegib. Tumor growth of the patients is represented in Figure 4.

**Figure 4.**
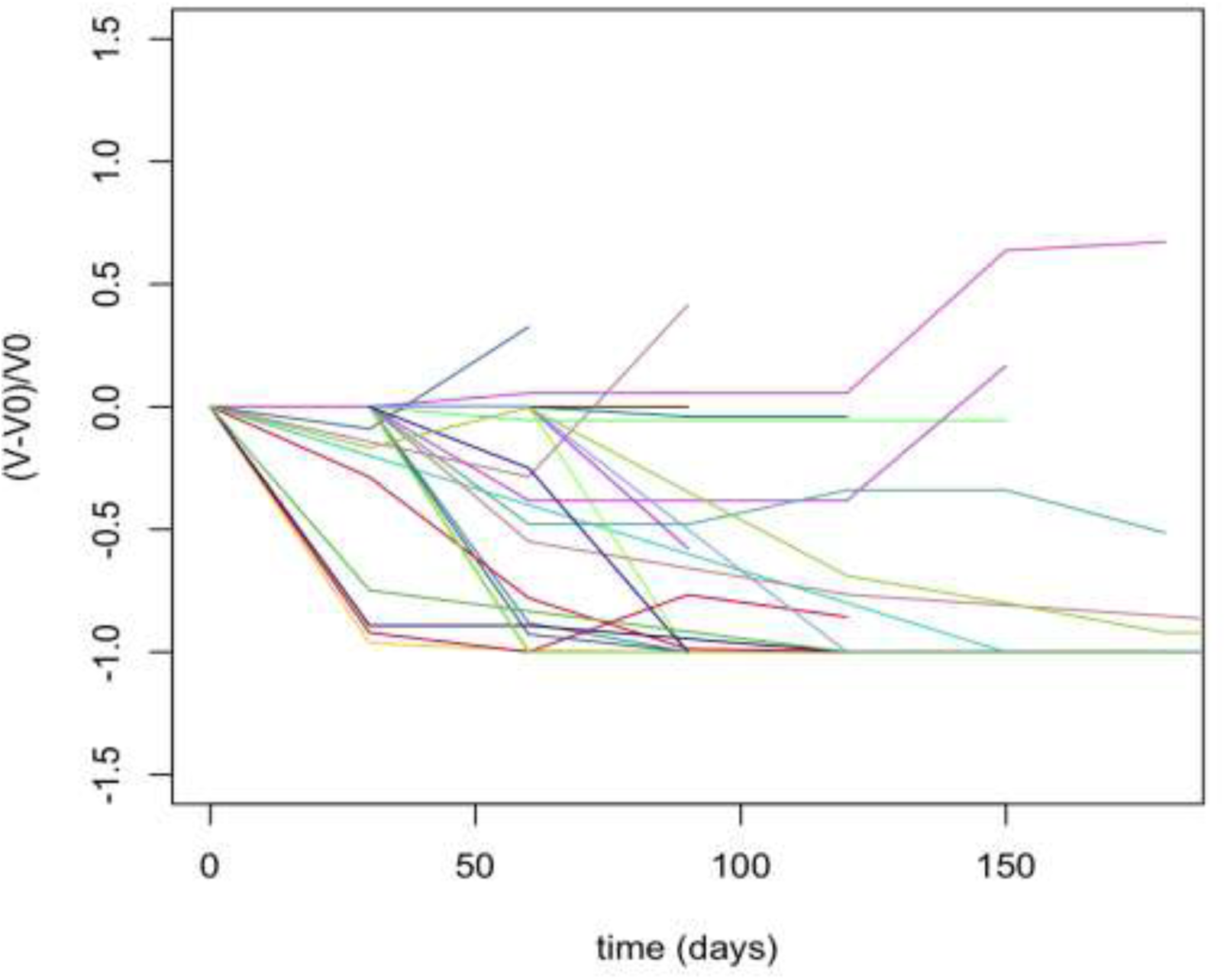
Tumor growth of the patients of the OPTIVISMO study. The tumor growth was calculated as: (V-V0) / V0, where V denoted the last estimated tumor volume and V0 the first available volume (n=22); *5 patients were excluded of the analysis because of missing values on tumor size*.

### 3.8 Correlation tests

The correlation between the concentration of AAG and the trough plasma concentrations of vismodegib was statistically significant (Spearman’s ρ = 0.6733, p-value =1.662e-12) (Figure 5), which is consistent with the high and saturable binding of vismodegib to AAG. The tumor volume slope was significantly correlated to the mean concentration of vismodegib (Spearman’s ρ = 0.4274, p-value = 0.02941) (Figure 6).

**Figure 5.**
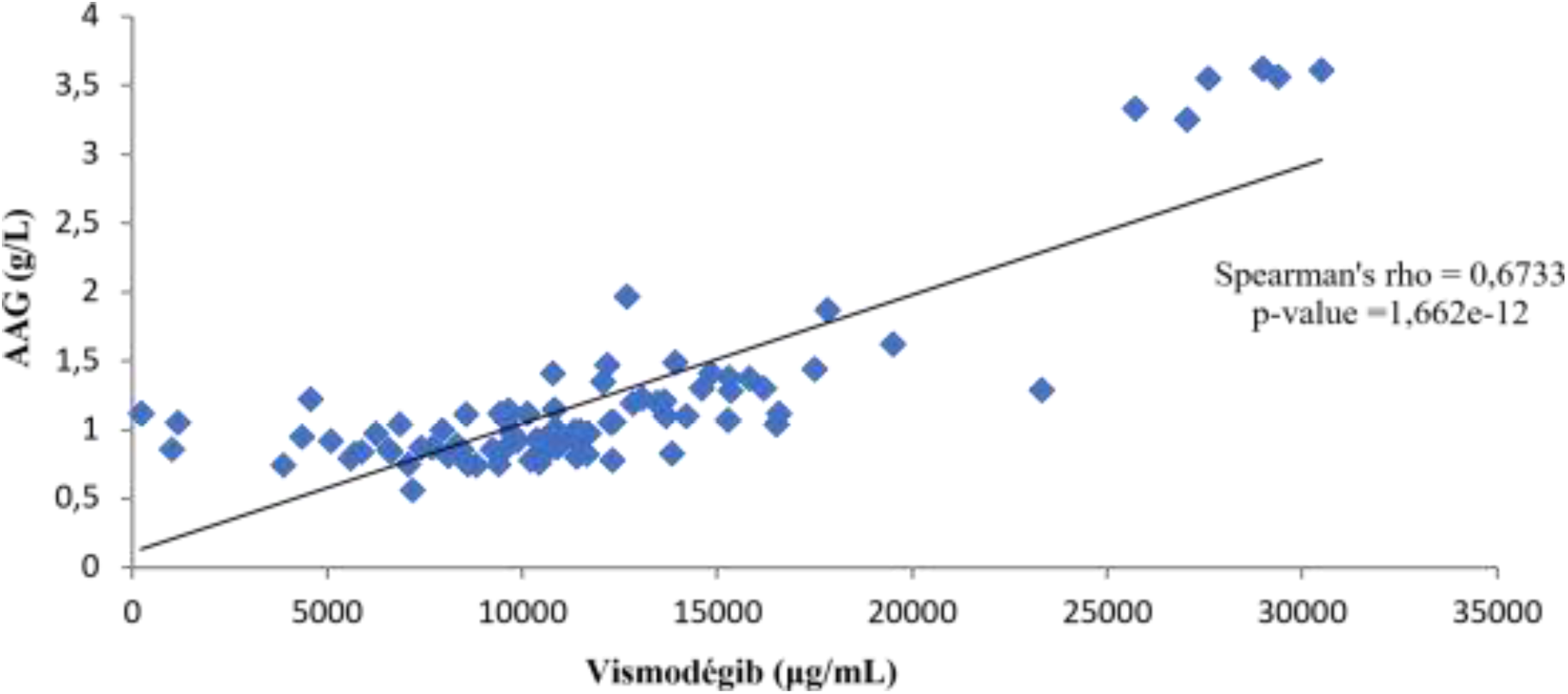
Correlation between total vismodegib concentrations and acid alpha-1-glycroprotein (AAG) concentration.

**Figure 6.**
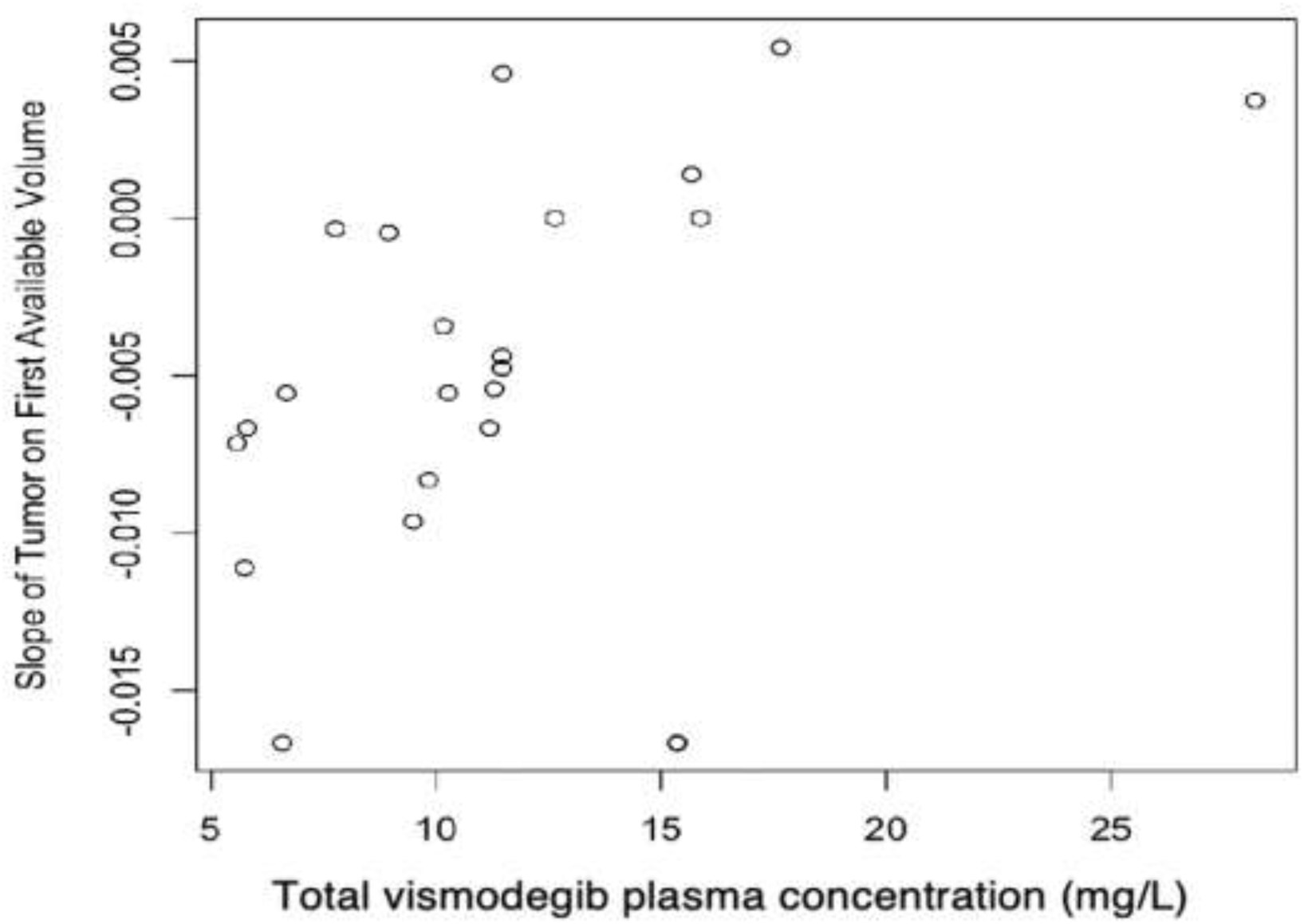
Correlation between mean of total vismodegib concentration (per patient) and the estimated volume tumor slope (n=22) (Spearman’s ρ = 0.4274, p-value =0.02941). *5 patients were excluded of the analysis because of missing values on tumor size*.

### 3.9 Discussion

In our study, we obtained pharmacodynamic and pharmacokinetic data using vismodegib in a real-life clinical practice. Total vismodegib plasma concentrations observed were similar to those found in the literature [12]. However, analysis of patients included in the OPTIVISMO study revealed an older population with more comorbidities compared to patients included in the drug’s clinical development studies [12,13]. The most common AEs were muscle spasm, dysgeusia, and alopecia, which appeared to be common effects of inhibitor of the Hedgehog signaling pathway class [14].

The pharmacokinetic analysis confirmed previously published data in clinical trials, as a high inter-individual variability of concentrations, and a high correlation with AAG levels [4]. The overall low intra-patient variability between one and 6 months of follow-up indicated that one blood sample, drawn when the PK steady-state is reached, is likely to be sufficiently informative in order to estimate the systemic exposure of a patient to vismodegib. In case of significant changes in AAG level, one should expect a significant impact on vismodegib concentration for a patient.

One limitation of our study is that only univariate statistical analyses have been performed, so that further multivariate analysis are needed to confirm that AAG is the only covariate influencing vismodegib concentrations. For the PK/PD assessment, we notably observed that patients with lower plasma exposure were those who experienced a better response under vismodegib according to RECIST criteria and regarding tumor growth evolution. One hypothesis is that high AAG levels binding vismodegib with high affinity could be a factor limiting the drug’s tissue distribution. However, the groups were not homogeneous according to other variables as renal function, age and weight, potentially influencing vismodegib concentrations, so that a further statistical analysis should be performed to assess whether vismodegib concentration is the main independent factor associated with response.

Our results highlighted the need of analysing unbound vismodegib concentrations to provide a better understanding of the PK/PD relationship of vismodegib described in this study. To date, there are very few published data on the free form of vismodegib in the literature. Our study may provide free-form data on a larger cohort (n=25). In a study by Deng et al. the free fraction was measured in only three patients by rapid equilibrium dialysis (RED) followed by HPLC-MS/MS analysis [15]. To experimentally determine a percentage of protein binding, the separation of unbound and bound forms can be achieved by various techniques, including ultrafiltration, ultracentrifugation, gel filtration, capillary electrophoresis, and equilibrium dialysis. One perspective on this study would be to develop a method for separating free and bound forms of vismodegib by equilibrium dialysis, which reproduces the binding of a drug to plasma proteins and the equilibrium reached under physiological conditions [16]. Finally, modelling BCC growth by mathematical equations and determining the individual parameters that have an impact on this growth are likely to be one another working perspective.

## 5 WHAT IS NEW AND CONCLUSION

Our study explores the PK/PD relationship of vismodegib in patients with BCC. This is the first study which reported PK data obtained in patients treated with vismodegib in routine clinical practice, as previous data and PK models have been obtained from randomized studies. Our study confirmed the strong influence of AAG levels on vismodegib protein binding and thus on plasma concentrations. The PK/PD correlations showed that patients with the lowest plasma concentrations respond best to treatment. The mathematical estimation of tumor volume revealed that tumor globally decreased in our population of patients with BCC during the follow-up and that growth was correlated with vismodegib PK, which was in line with the correlation observed with efficacy/safety data.

## Data Availability

All data produced in the present study are available upon reasonable request to the authors

